# Early Development of Selective Motor Control in Preterm Infants With and Without Cerebral Palsy

**DOI:** 10.1101/2025.05.09.25327335

**Authors:** Colleen Peyton, Theresa Sukal Moulton, David Aaby, Ryan Millman, Raye Ann deRegnier, Arend Bos, Jules Dewald

**Affiliations:** Department of Physical Therapy and Human Movement Sciences, Feinberg School of Medicine, Northwestern University, Chicago, IL, USA; Department of Pediatrics, Feinberg School of Medicine, Northwestern University, Chicago, IL, USA; Department of Preventive Medicine, Feinberg School of Medicine, Northwestern University, Chicago, IL, USA; University of Illinois at Chicago, Chicago, IL USA; Ann and Robert H. Lurie Children’s Hospital, Chicago, IL, USA, Division of Neonatology, Department of Pediatrics, Beatrix Children’s Hospital; University Medical Center Groningen, University of Groningen, Groningen, the Netherlands; Department of Biomedical Engineering, McCormick School of Engineering, Northwestern University, Evanston, IL USA

**Author notes:** Corresponding Author: Colleen Peyton. 645 N. Michigan Ave, Suite 1100, Chicago IL 60611.

## Abstract

**Objectives:** To characterize early developmental trajectories of selective motor control (SMC) in very preterm infants and examine associations with later cerebral palsy (CP) diagnosis and gross motor function.

**Methods:** Very preterm infants (<32 weeks’ gestation) were recorded every 2–4 weeks until 5 months post-term age (PTA). SMC was scored from 352 videos (n=47 infants; 12 with CP) using BabyOSCAR, a validated observational tool. Linear mixed models examined SMC trajectories by CP diagnosis and Gross Motor Function Classification System (GMFCS) level. ROC curves tested the ability of early SMC change (40–45 weeks) to predict CP.

**Results:** SMC scores increased over time, but infants with CP showed slower gains. Between 41–63 weeks, group differences emerged and widened (p<0.001). Change in BabyOSCAR score from 40–45 weeks predicted CP with 92% sensitivity and 100% specificity (AUC=0.98). GMFCS groups showed distinct trajectories, with children classified as GMFCS III–V changing scores less. Infants with unilateral CP showed increasing asymmetry from 42 weeks PTA.

**Conclusions:** SMC develops rapidly after term age but is altered in infants with CP, particularly among those later classified as GMFCS III–V. Early trajectories may reflect emerging corticospinal connectivity and offer a clinically useful marker of functional motor outcomes.

---

Selective motor control (SMC), he ability to move one joint independently of others, is essential for skilled movement^1^ but remains poorly understood in infancy. While SMC is the greatest determinant of functional ability in cerebral palsy (CP)^2^, its emergence and trajectory have not been directly studied. It is thought to arise from targeted monosynaptic connections between the developing corticospinal tract (CST) and specific motor neuron pools^3^. Investigating how SMC develops could provide insight into the maturation of motor pathways and inform early identification of movement disorders.

The first fetal movements appear by 7.5 weeks post-menstrual age (PMA)^4^ and continue throughout fetal development. Initially, fetal movement is believed to be primarily driven by brainstem and spinal neuronal networks^5–9^. Whole body movements from 9.5 weeks PMA^10^ support large-scale neuronal networks^6,8,11,12^ Transient cortical structures like the subplate facilitate early sensorimotor processing before thalamocortical and corticospinal circuits emerge^12,13^. With advancing gestation, these transient circuits are gradually pruned and replaced by more permanent cortical pathways^13–15^. By term age, rapid structural changes—including synaptogenesis, myelination, and somatotopic refinement^13,14^—support the increasing involvement of cortical systems in motor control^16^. This transition may underlie SMC emergence. However, if cortical activity is disrupted during early development, as may occur in infants with CP, brainstem motor circuits may maintain or even expand their influence over movement, reflecting a form of competition between descending motor pathways^9,16,17^.

Studying motor development in human fetuses is challenging, but very preterm infants (<32 weeks PMA) provide a unique opportunity to observe motor behavior at an age when they would typically still be developing in utero. Their movement reflects neural circuitry at that gestational age, offering insight into early motor control. However, when SMC first emerges in infancy and how it evolves in relation to cortical development remain unknown.

To explore this question, we utilized BabyOSCAR^18^, a non-invasive observational tool designed to quantify spontaneous SMC in infancy. By systematically measuring SMC across early development, BabyOSCAR provides a unique opportunity to assess how SMC emerges over time in preterm infants. This study aimed to characterize the early trajectory of SMC from preterm age to 5 months PTA in very preterm infants with and without CP. We hypothesized that SMC would increase over time in all infants, but those later diagnosed with CP will exhibit significantly lower SMC by 3 months PTA compared to their peers.

## Method

### Participants

Very preterm infants (<32 weeks GA) were recruited prospectively from two neonatal intensive care units (NICU). Infants were then followed until ≥18 months of age to determine if they received an eventual diagnosis of CP. CP diagnoses were made by trained physicians at ≥18 months. Gross Motor Functional Classification System (GMFCS), and body topography (unilateral or bilateral) level were recorded for children with CP. Guardians provided written consent for video use in research.

### Video recordings

Infants were video recorded every 2-4 weeks from enrollment to 5 months PTA, in the NICU or after discharge at home, clinic, or lab. Videos (3-10 min) captured spontaneous movement during a calm, alert state without a pacifier. Before 40 weeks PTA, infants were recorded asleep or awake; after 40 weeks, only during calm, alert states. All limbs were visible for scoring.

### BabyOSCAR

BabyOSCAR quantifies isolated joint motion—movement without associated intra-limb or mirrored motion^18^. Each isolated movement is scored as 1. There are 18 joint motions scored for the arms (arm score) and 14 for the legs (leg score). Arm and leg scores sum to a total BabyOSCAR score (range 0 to 32)^18^. An asymmetry score is calculated by subtracting scores on the left side of the body (sum of left arm and leg) from the right side of the body (sum of right arm and leg)^18^. Videos were rated by two raters (CP, TM).

## Statistical Analysis

All analyses were performed using R version 4.1.2 (R Foundation for Statistical Computing, Vienna, Austria). Sample characteristics were summarized as either n (%) or as median (intraquartile range; IQR), as appropriate. We examined: (1) total, (2) arm, (3) leg, and (4) asymmetry BabyOSCAR scores, with higher values denoting greater asymmetry. We used linear mixed-effects regression models to examine (1) changes in scores as infants age (2) differences in the rate of change in scores among those infants with CP and those without CP. Random intercepts and random slopes for age were included for each infant to account for the variability in motor development trajectories within and across infants. Age was modeled using restricted cubic splines (4 knots) to account for non-linear relationships over time. As we hypothesized that the association between age and BabyOSCAR would differ by CP diagnosis, we included age*CP interaction terms in the models. We used likelihood ratio tests (LRT) to determine significance of the interaction terms. We generated marginal BabyOSCAR score estimates over time by CP diagnosis. We repeated these analyses, further categorizing CP by GMFCS level (no CP, GMFCS I-II, GMFCS III-V)

We generated a ROC curve to evaluate BabyOSCAR score change (Δ40–45 weeks PMA) in predicting CP. This window was chosen because it captures the onset of functional corticospinal influence, when group differences in SMC first appear. We fit a linear mixed effect model, including a fixed effect for age, and random intercepts and slopes for each infant. Infant-specific score slopes (change from 40–45 weeks PMA) were extracted. The optimal cutpoint was determined using the Youden index. A p-value of ≤0.05 was considered statistically significant.

## Results

Forty-seven infants and 352 total video recordings scored with the BabyOSCAR were included in the analysis, with a median of 9 longitudinal video recording per infant (IQR: 5,11). Twelve infants (26%) were later diagnosed with CP by 18 months of age (Table 1). All 6 children with unilateral CP were later characterized as having GMFCS I-II, while all the 6 children with bilateral CP all were classified as GMFCS III-V.

**Table 1.** Subject characteristics of 47 children and cerebral palsy specific characteristics among 12 children.

| Subject Characteristic |  | N=47<sup>a</sup> |
| --- | --- | --- |
| Gestational age (weeks) |  | 28 (27, 30) |
| Birthweight (g) |  | 1080 (830, 1430) |
| Female |  | 17 (36) |
| Race |  |  |
|  | Asian | 4 (10) |
|  | Black or African American | 2 (4) |
|  | White | 35 (74) |
|  | Unknown | 6 (13) |
| Ethnicity |  |  |
|  | Hispanic or Latino | 4 (9) |
|  | Not Hispanic or Latino | 32 (68) |
|  | Unknown | 11 (23) |
| Multiple gestation birth |  | 20 (42) |
| Intraventricular hemorrhage |  | 19 (40) |
| Periventricular leukomalacia |  | 4 (9) |
| Ventriculoperitoneal shunt |  | 3 (6) |
| Bronchopulmonary dysplasia<sup>b</sup> |  | 26 (55) |
| Necrotizing enterocolitis<sup>c</sup> |  | 2 (4) |
| Treated retinopathy of prematurity<sup>d</sup> |  | 7 (16) |
| Diagnosis of cerebral palsy |  | 12 (26) |
| Cerebral Palsy Characteristics |  | N=12<sup>a</sup> |
| Topography |  |  |
|  | Bilateral | 6 (50) |
|  | Unilateral | 6 (50) |
| Unilateral paretic side |  |  |
|  | Right | 3 (50) |
|  | Left | 3 (50) |
| Type of cerebral palsy |  |  |
|  | Spastic | 9 (75) |
|  | Dyskinetic | 1 (8) |
|  | Mixed | 1 (8) |
|  | Unknown | 1 (8) |
| GMFCS<sup>e</sup> level |  |  |
|  | I | 4 (33) |
|  | II | 2 (17) |
|  | III | 2 (17) |
|  | IV | 2 (17) |
|  | V | 2 (17) |

<sup>a</sup>Median (IQR); n (%), <sup>b</sup>Required supplemental oxygen $\geq 35$ weeks gestation, <sup>c</sup>Treated with drain or laparoscopy, <sup>d</sup>Treated with Avastin and/or laser therapy, <sup>e</sup>GMFCS: Gross Motor Functional Classification System; measured $\geq 18$ months corrected age

### Trajectories of Total BabyOSCAR score over time

Figure 1 shows a spaghetti plot of each infant’s A) Total, B) Arm, and C) Leg BabyOSCAR scores over time with the overall model estimates over time by CP diagnosis in children with and without CP.

**Figure 1.**
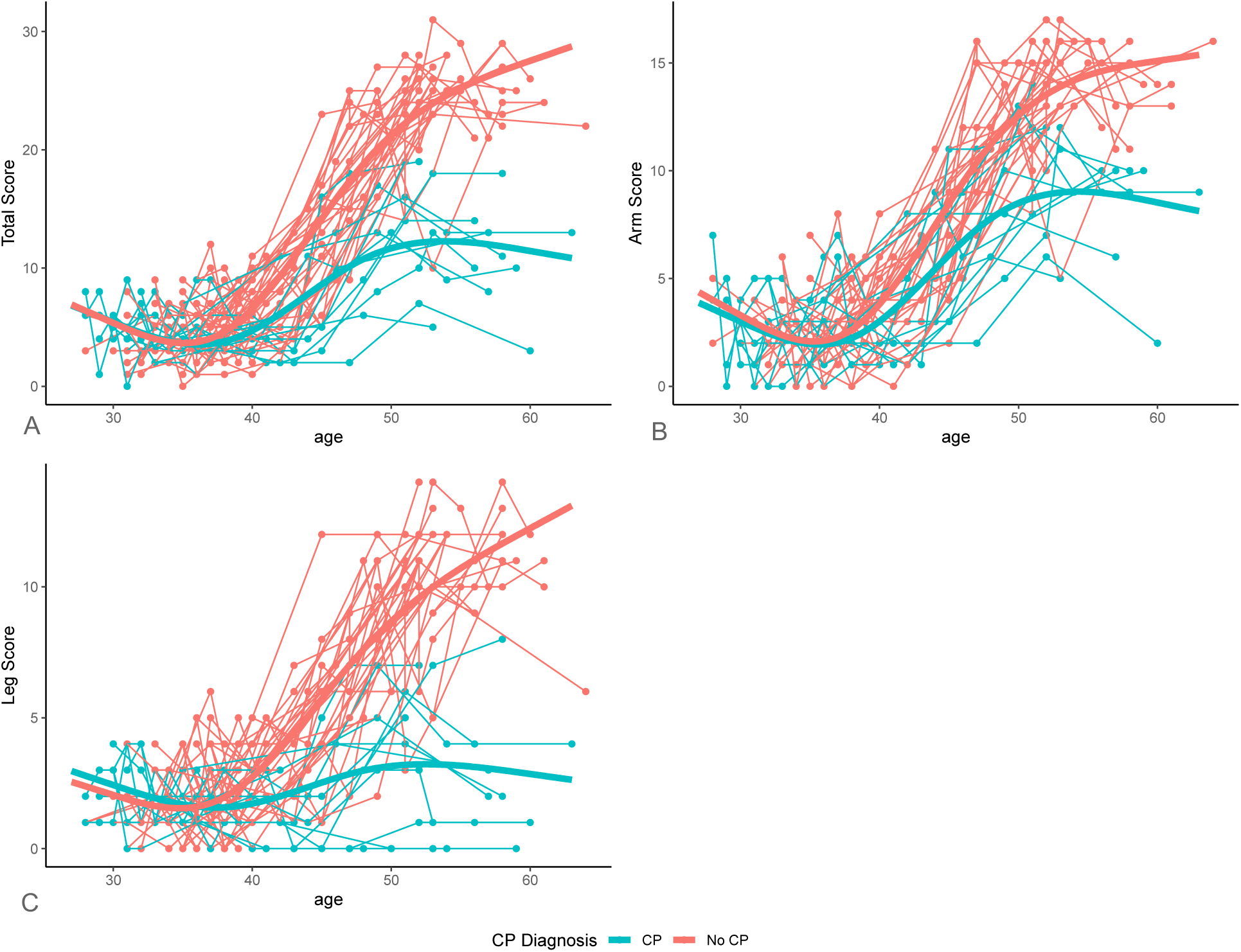
Spaghetti plot of A) Total BabyOSCAR Score B) BabyOSCAR Arm Score C) BabyOSCAR Leg Score by Post Menstrual Age in weeks in Children with and Without CP Dots: individual BabyOSCAR scores; thin lines: individual participant trajectories over time; solid lines: Loess-smoothed trends for each group

We found a significant difference in Total BabyOSCAR trends over time between infants with and without CP (p<0.001). Although both groups showed increases in Total BabyOSCAR scores over time, the rate of improvement was slower in infants with CP. Total BabyOSCAR scores did not differ significantly between infants with and without CP between 27-40 weeks PMA (p>0.05, Supplemental Table 1). However, by 41 weeks PMA, infants with CP scored lower than peers without CP (estimate=-2.5, p=0.012, Supplemental Table 1), with progressively larger differences from 42 weeks and older (p<0.001, Supplemental Table 1).

Notably, we also found a significant interaction between CP diagnosis and age (p=0.042; mixed model analysis), indicating that although both groups showed increases in BabyOSCAR scores over time, the rate of improvement was slower in infants with CP. As development progressed, the gap widened, with infants with CP exhibiting increasingly lower scores compared to their peers without CP (3.2 points lower at 42 weeks, p<0.001; 18.4 points lower at 63 weeks, p<0.001, Supplemental Table 1).

### Trajectories of BabyOSCAR Arm and Leg scores over time

Arm and Leg trajectories also differed significantly over time between infants with and without CP (Likelihood ratio test for interaction model: Arm χ²=30.6, df=3, p<0.001; Leg χ²=50.3, df=3, p<0.001). No significant group differences were observed in either subscale before 41 weeks PMA (p>0.05 Supplemental Tables 2 and 3); however Leg scores began to diverge significantly one week earlier than Arm scores. The first significant group difference in Leg scores appeared at 41 weeks, with lower scores in the CP group compared to those without CP (estimate=-1.4, p=0.004; Supplemental Table 3), followed by a difference in Arm scores at 42 weeks (estimate=-1.4, p=0.024; Supplemental Table 2). Both subscales showed progressively larger differences after 42 weeks (p<0.001; Supplemental Tables 2 and 3).

### Predictive model of Total BabyOSCAR score over time

Figure 2 displays the receiver operating characteristic (ROC) curve, illustrating the performance of using the change in BabyOSCAR score between 40-45 weeks PMA to predict CP diagnosis ≥18 months of age. The change in score showed excellent predictive performance (AUC = 0.98; Figure 2). The optimal cutpoint was 2.9 over the 5-week period, with 92% sensitivity and 100% specificity, as well as 100% positive predictive value and 97% negative predictive value. That is, if an infant does not see an increase of ≥3 points on the BabyOSCAR, then the infant is likely to have CP when evaluated ≥18 months of age. These findings suggest that minimal SMC improvement across this 5-week window may warrant closer clinical monitoring and developmental follow-up.

**Figure 2.**
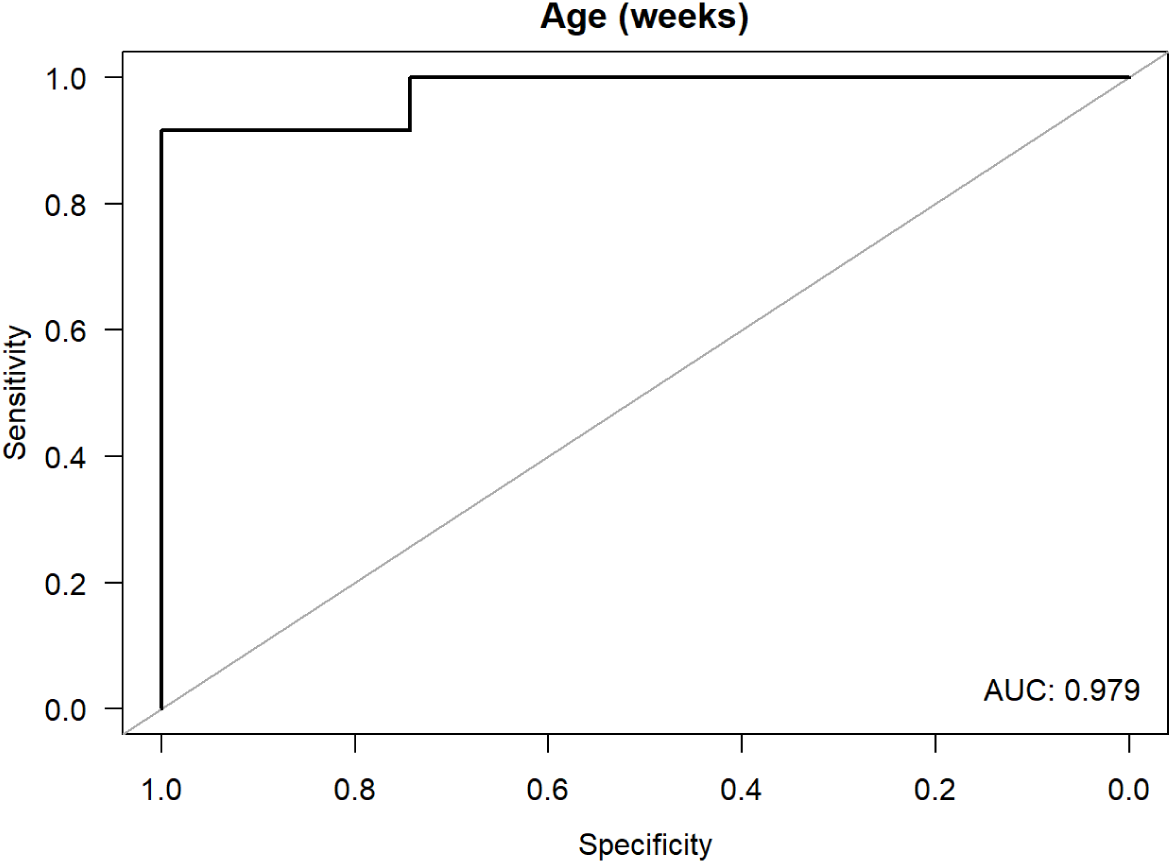
Receiver Operating Characteristic Curve for change in BabyOSCAR score between 40-45 weeks PMA in prediction of cerebral palsy

### Trajectories of Total BabyOSCAR score over time by Gross Motor Functional Classification System outcome (GMFCS)

We examined Total BabyOSCAR score trajectories across three groups: children without CP (n=35), children with GMFCS I–II (n=6), and children with GMFCS III–V (n=6). Trajectories differed significantly across groups (Likelihood ratio test for interaction model: χ²=15.7, df=3, *p*=0.001). Scores increased most in children without CP, less in GMFCS I-II, and least in GMFCS III-V (Figure 3). Differences between GMFCS subgroups appeared later in development: at 49 weeks PMA, infants with GMFCS III–V had significantly lower scores compared to those with GMFCS I–II (estimate=-4.6, p=0.04, Supplemental Table 4), with differences increasing progressively through 59 weeks PMA (estimate=-7.4, p=0.04; Supplemental Table 4).

**Figure 3.**
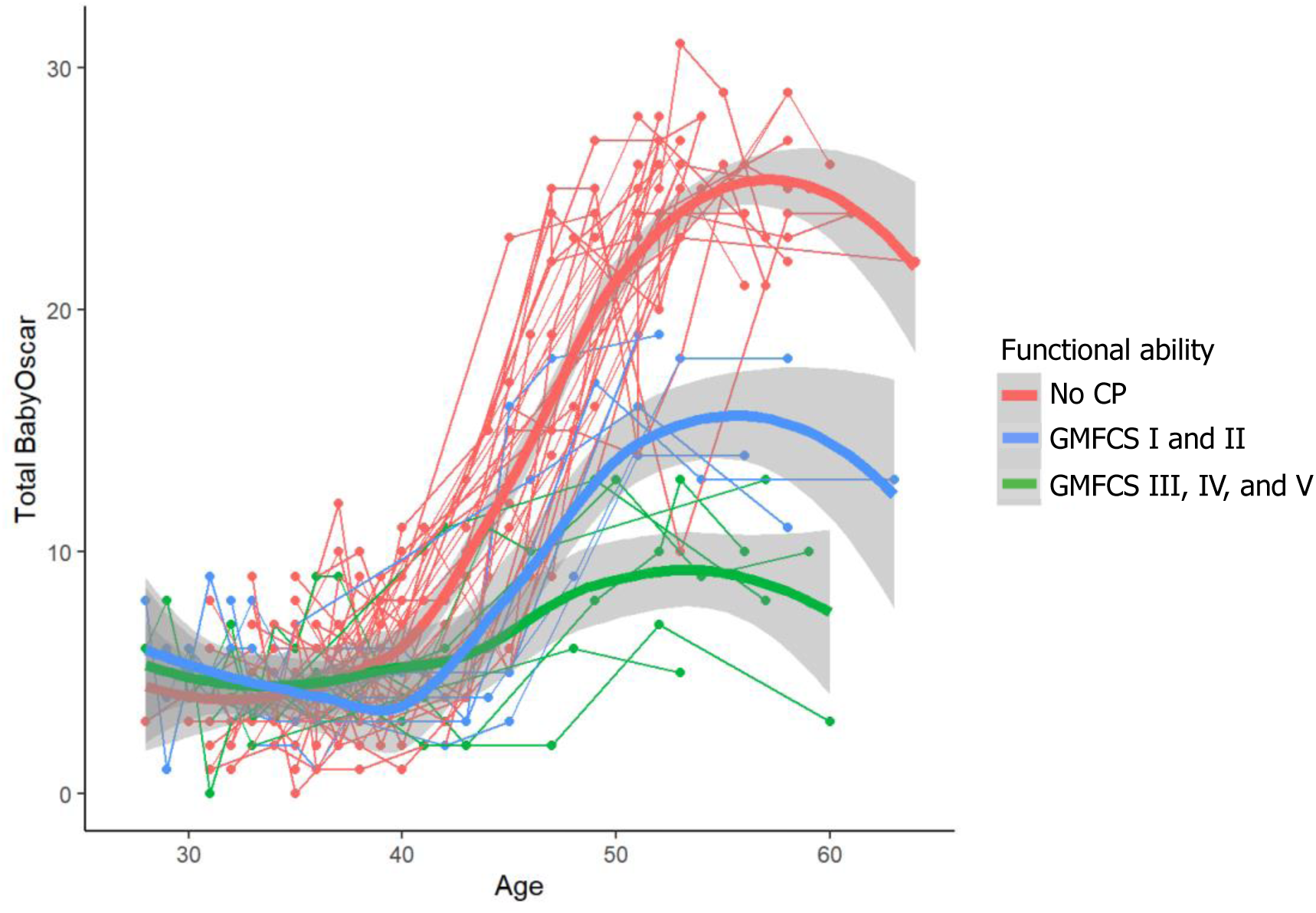
Spaghetti plot of Total BabyOSCAR Score among children without CP, children with GMFCS I-II and children with GMFCS III-V

### Trajectories of Total BabyOSCAR score over time by CP body distribution including children with bilateral CP, children with unilateral CP and children without CP

We examined BabyOSCAR asymmetry scores (absolute values) over time in three groups: children without CP, children with bilateral CP and children with unilateral CP. Trajectories differed significantly by topography (Likelihood ratio test for interaction model: χ²=20.0, df=3, *p*<0.001), where infants with unilateral CP had a distinct trajectory from children with bilateral CP or no CP. During the preterm period, there were no significant differences among asymmetry scores between the three groups. However, at 42 weeks PMA, the first significant difference was seen between infants with unilateral CP and infants without CP where at 42 weeks, infants with unilateral CP began having a significantly higher asymmetry score than infants without CP (estimate=0.9, p=0.029; Supplemental Table 5). This difference increased with increasing gestation to a difference of 5.6 higher asymmetry score in the unilateral CP by 63 weeks PMA (p<0.001). Infants with unilateral CP also showed significantly higher asymmetry scores than those with bilateral CP starting at 44 weeks (estimate =1.3, p=0.018, Supplemental Table 6), with increasing asymmetry scores over time to an increased asymmetry score of 4.6 by 63 weeks PMA (p=0.012, Supplemental Table 6). There were no differences over time in asymmetry scores between infants with bilateral CP and infants without CP.

## Discussion

Early SMC emerges differently in infants with and without CP, with group differences first evident around 41 weeks gestational age and continuing to widen over the first 4 months after term age. This period aligns with the developmental window in which human corticospinal and corticostriatal circuits are becoming increasingly active and organized^19–21^, suggesting the increasing role of the descending cortical pathways as neural substrates for independent joint motion, a capacity that is reduced or altered in infants with CP. Notably, increased SMC capacity in the first month predicted later CP diagnosis.

Although all infants later diagnosed with CP sustained their brain injuries prior to 32 weeks gestational age—typically within the first 72 hours of life—we observed no significant group differences in SMC during the preterm period (Fig 1). This finding suggests that early spontaneous movement in fetal and preterm development is largely mediated by bulbospinal pathways, which drive motor output before the corticospinal tract contributes functionally to support SMC. Motor activity is primarily driven by subcortical systems early in gestation, with increasing cortical contributions later^16,22,23,24^ and neurophysiological studies demonstrate that corticospinal projections reach the spinal cord by the late second trimester^19^. However, their functional efficacy increases substantially after term^25^, driven by synaptic reorganization, dendritic arborization, and myelination^12,14^. The resolution of subplate structures and concurrent emergence of direct corticospinal influence over spinal motor neurons may support the initial capacity for independent joint motion, as measured by BabyOSCAR. Therefore, injury may not manifest behaviorally until cortical influence becomes critical for isolated movements.

Previously, BabyOSCAR was validated only in infants with spastic CP^18^, but this prospective study included two children with dyskinetic or mixed-type CP. Despite differences in CP subtype, these infants followed a similar pattern, showing no early divergence in SMC during the preterm period, followed by slower post-term increases and lower overall SMC scores compared to infants without CP. Dyskinetic and mixed-type CP subtypes are often associated with injury to the basal ganglia and/or subthalamic nuclei^26^, suggesting that SMC may also depend on later-developing cortico-subcortical networks. Corticostriatal fibers peak by 29 weeks PMA, but full basal ganglia circuits form closer to term^21^. This timeline supports the possibility that basal ganglia contributions to SMC emerge later, once functional connections with cortical motor areas are fully established.

We observed slower increases in SMC and lower overall capacity for independent joint motion in infants with CP as compared to infants without CP (Figure 1), with the most pronounced differences among children with GMFCS III-V (Figure 3). These trajectory patterns likely reflect the extent and laterality of corticospinal disruption. In infants with GMFCS I-II (all with unilateral CP), residual corticospinal activity from the less affected hemisphere may support some preservation of independent joint control, consistent with prior findings in older children^27,28^. In contrast, infants with GMFCS III-V (all with bilateral CP) likely experienced more widespread disruption of cortical motor pathways, limiting the development of SMC. In these cases, descending brainstem pathways such as the vestibulospinal and reticulospinal tracts may exert greater influence. Animal models suggest that, when cortical activity is inhibited, brainstem pathways may maintain or expand their developmental influence^17,29^, potentially explaining the reduced SMC capacity we observed in these infants. We previously found that infants later classified as GMFCS levels III-V more frequently displayed coupled or mirrored limb movements by 3 months^18,30^, patterns consistent with bulbospinal projections, which synapse bilaterally and span multiple spinal segments^31^. These circuits may remain hyperexcitable, as seen in adults with spastic CP^32^.

Infants with unilateral CP in our study did not show differing asymmetry from infants without CP or children with bilateral CP during the preterm period (Figure 4). However, by approximately 5 weeks post-term age, infants with unilateral CP began to show significantly greater asymmetry with this difference continuing to widen over time. Similar findings have been reported previously by others, who observed the emergence of asymmetrical segmental movements, defined as distinct joint movements of the distal limbs, in infants later diagnosed with unilateral CP by 3 months of age in infants born preterm^33^ and 2 months of age in term-born cohorts^34^. The earlier asymmetry seen in our sample may be due to methodological differences: while all assessed spontaneous movement, BabyOSCAR captures capacity for isolated joint motion across both upper and lower extremities, incorporating both proximal and distal segments. Nevertheless, in all cases, the delayed emergence of asymmetry is consistent with the developmental window during which corticospinal projections transition from early bilateral innervation toward a predominantly contralateral organization^35^. In cases of unilateral CP, activity-dependent competition between hemispheres may favor the retention of ipsilateral projections from the non-lesioned hemisphere^29^—a mechanism that may manifest functionally as early as 2-4 weeks post-term. Notably, such asymmetries are unlikely to be observable during the preterm period, highlighting that early motor assessments conducted in the NICU may not yet capture behavioral signs that presage a diagnosis of unilateral CP. These findings support continued developmental monitoring after discharge.

**Figure 4.**
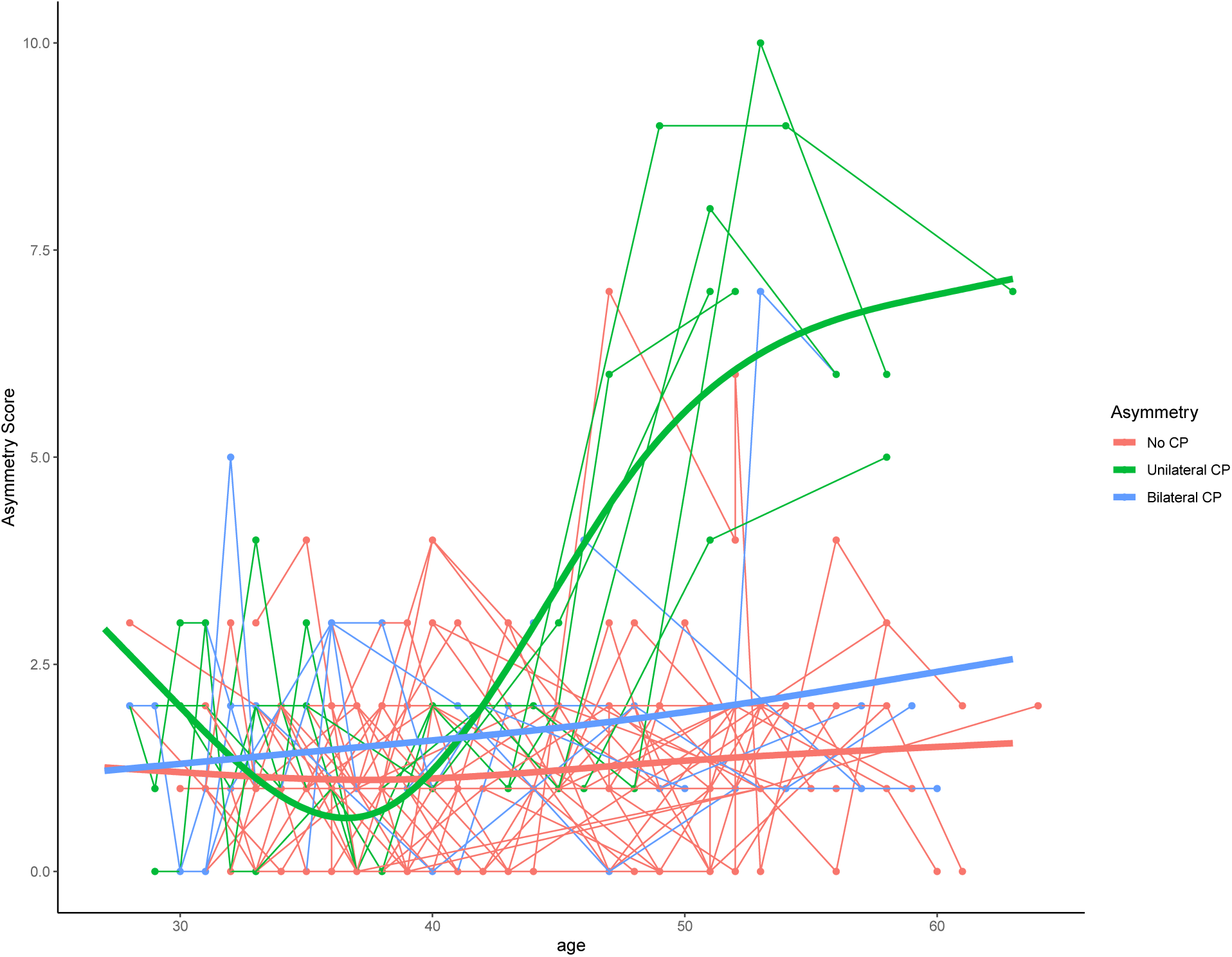
Spaghetti plot of BabyOSCAR Asymmetry Score (absolute value) among children without CP, children with unilateral CP and bilateral CP

We were surprised to find that the rate of change in total BabyOSCAR score between 40-45 weeks PMA was highly predictive of a later CP diagnosis (AUC 0.98), with excellent sensitivity and specificity. While we previously found that total BabyOSCAR score at 3 months of corrected age could predict later CP diagnosis, we did not anticipate that such a short and early developmental window would also yield strong prognostic information. This likely reflects the emergence of functional corticospinal influence during this window. If capacity of SMC fails to improve during this period, it may indicate a disruption of descending motor circuits. Continued use of BabyOSCAR beyond this window, especially up to 3 months corrected age, can provide additional prognostic clarity, including insights into body topography (via asymmetry scores) and later gross motor abilities (via GMFCS level), as shown in our earlier work. These results support SMC as a mechanistically grounded, developmentally sensitive target for early CP detection.

However, the major limitation of this study is the small number of subjects assessed. Recruiting preterm infants without known CP outcome is difficult and requires multicenter collaboration. A larger number of infants is needed to confirm these findings and hypotheses. Furthermore, we have studied very preterm infants in this study to understand the ability to generate SMC in the preterm period. However, additional studies, with infants born at term, would be useful to generalize findings to a broader population of infants.

Early differences in SMC between infants with and without CP appear shortly after term age, reflecting the functional emergence of descending motor circuits during this critical developmental window. BabyOSCAR is a developmentally grounded, scalable tool that may help clinicians identify infants who show early signs of altered motor development and benefit from timely referral or intervention. These findings underscore the value of monitoring SMC trajectories after NICU discharge to inform early clinical decision-making; however, larger studies across diverse populations are needed to validate the tool’s predictive utility and support broader implementation.

## Data Availability

All data produced in the present study are available upon reasonable request to the authors

---

**Supplemental Table 1.** Model estimates of Total BabyOSCAR score differences in children with CP relative to children without CP, by gestational age (weeks)

| Gestational age (weeks) | Estimate | 95% Confidence intervals | p-value |
| --- | --- | --- | --- |
| 27 | 0.2 | (-3.2, 3.6) | 1 |
| 28 | 0.2 | (-2.7, 3.2) | 1 |
| 29 | 0.2 | (-2.3, 2.8) | 1 |
| 30 | 0.3 | (-1.8, 2.4) | 0.999 |
| 31 | 0.3 | (-1.5, 2.1) | 0.996 |
| 32 | 0.3 | (-1.2, 1.8) | 0.991 |
| 33 | 0.3 | (-1, 1.6) | 0.989 |
| 34 | 0.2 | (-1, 1.5) | 0.995 |
| 35 | 0.1 | (-1.1, 1.4) | 1 |
| 36 | -0.1 | (-1.4, 1.3) | 1 |
| 37 | -0.3 | (-1.8, 1.1) | 0.987 |
| 38 | -0.7 | (-2.2, 0.8) | 0.861 |
| 39 | -1.2 | (-2.7, 0.3) | 0.499 |
| 40 | -1.8 | (-3.3, -0.2) | 0.134 |
| 41 | -2.4 | (-3.9, -0.9) | 0.013 |
| 42 | -3.1 | (-4.6, -1.6) | <0.001 |
| 43 | -3.9 | (-5.4, -2.4) | <0.001 |
| 44 | -4.7 | (-6.3, -3.1) | <0.001 |
| 45 | -5.5 | (-7.2, -3.8) | <0.001 |
| 46 | -6.4 | (-8.2, -4.6) | <0.001 |
| 47 | -7.2 | (-9.2, -5.3) | <0.001 |
| 48 | -8.1 | (-10.1, -6) | <0.001 |
| 49 | -8.9 | (-10.9, -6.8) | <0.001 |
| 50 | -9.6 | (-11.7, -7.6) | <0.001 |
| 51 | -10.4 | (-12.5, -8.3) | <0.001 |
| 52 | -11.1 | (-13.3, -9) | <0.001 |
| 53 | -11.8 | (-14, -9.7) | <0.001 |
| 54 | -12.5 | (-14.8, -10.3) | <0.001 |
| 55 | -13.2 | (-15.6, -10.9) | <0.001 |
| 56 | -13.9 | (-16.5, -11.3) | <0.001 |
| 57 | -14.6 | (-17.4, -11.7) | <0.001 |
| 58 | -15.2 | (-18.4, -12) | <0.001 |
| 59 | -15.9 | (-19.4, -12.3) | <0.001 |
| 60 | -16.5 | (-20.5, -12.6) | <0.001 |
| 61 | -17.2 | (-21.5, -12.8) | <0.001 |
| 62 | -17.8 | (-22.6, -13) | <0.001 |
| 63 | -18.4 | (-23.7, -13.2) | <0.001 |

**Supplemental Table 2.** Model estimates of BabyOSCAR Arm score differences in children with CP relative to children without CP, by gestational age (weeks)

| **Gestational age (weeks)** | **Estimate** | **95% Confidence intervals** | **p-value** |
| --- | --- | --- | --- |
| 27 | -0.5 | (-2.6, 1.6) | 0.989 |
| 28 | -0.4 | (-2.2, 1.4) | 0.99 |
| 29 | -0.4 | (-1.9, 1.2) | 0.99 |
| 30 | -0.3 | (-1.6, 1) | 0.991 |
| 31 | -0.2 | (-1.3, 0.9) | 0.993 |
| 32 | -0.2 | (-1.1, 0.7) | 0.995 |
| 33 | -0.1 | (-0.9, 0.7) | 0.997 |
| 34 | -0.1 | (-0.9, 0.6) | 0.998 |
| 35 | -0.1 | (-0.9, 0.6) | 0.997 |
| 36 | -0.2 | (-1, 0.7) | 0.993 |
| 37 | -0.3 | (-1.1, 0.6) | 0.976 |
| 38 | -0.4 | (-1.3, 0.5) | 0.912 |
| 39 | -0.6 | (-1.5, 0.4) | 0.726 |
| 40 | -0.8 | (-1.7, 0.1) | 0.405 |
| 41 | -1.1 | (-2, -0.2) | 0.13 |
| 42 | -1.4 | (-2.3, -0.5) | 0.024 |
| 43 | -1.7 | (-2.6, -0.8) | 0.003 |
| 44 | -2.0 | (-3, -1.1) | <0.001 |
| 45 | -2.4 | (-3.4, -1.4) | <0.001 |
| 46 | -2.7 | (-3.8, -1.6) | <0.001 |
| 47 | -3.1 | (-4.3, -1.9) | <0.001 |
| 48 | -3.4 | (-4.6, -2.2) | <0.001 |
| 49 | -3.8 | (-5, -2.5) | <0.001 |
| 50 | -4.1 | (-5.3, -2.8) | <0.001 |
| 51 | -4.4 | (-5.6, -3.1) | <0.001 |
| 52 | -4.6 | (-5.9, -3.4) | <0.001 |
| 53 | -4.9 | (-6.2, -3.6) | <0.001 |
| 54 | -5.2 | (-6.5, -3.8) | <0.001 |
| 55 | -5.4 | (-6.8, -4) | <0.001 |
| 56 | -5.7 | (-7.2, -4.1) | <0.001 |
| 57 | -5.9 | (-7.6, -4.2) | <0.001 |
| 58 | -6.1 | (-8, -4.2) | <0.001 |
| 59 | -6.3 | (-8.5, -4.2) | <0.001 |
| 60 | -6.6 | (-8.9, -4.2) | <0.001 |
| 61 | -6.8 | (-9.4, -4.1) | <0.001 |
| 62 | -7.0 | (-9.9, -4.1) | <0.001 |
| 63 | -7.2 | (-10.5, -4) | <0.001 |

**Supplemental Table 3.** Model estimates of BabyOSCAR Leg score differences in children with CP relative to children without CP, by gestational age (weeks)

| Gestational age (weeks) | Estimate | 95% Confidence intervals | p-value |
| --- | --- | --- | --- |
| 27 | 0.4 | (-1.2, 2.1) | 0.985 |
| 28 | 0.4 | (-1, 1.8) | 0.978 |
| 29 | 0.4 | (-0.8, 1.6) | 0.965 |
| 30 | 0.4 | (-0.6, 1.4) | 0.941 |
| 31 | 0.4 | (-0.5, 1.2) | 0.902 |
| 32 | 0.3 | (-0.4, 1) | 0.861 |
| 33 | 0.3 | (-0.3, 0.9) | 0.86 |
| 34 | 0.2 | (-0.4, 0.8) | 0.928 |
| 35 | 0.1 | (-0.5, 0.8) | 0.99 |
| 36 | 0.0 | (-0.7, 0.7) | 1 |
| 37 | -0.2 | (-0.9, 0.6) | 0.99 |
| 38 | -0.4 | (-1.2, 0.4) | 0.829 |
| 39 | -0.7 | (-1.5, 0.1) | 0.397 |
| 40 | -1.0 | (-1.8, -0.2) | 0.073 |
| 41 | -1.4 | (-2.2, -0.6) | 0.004 |
| 42 | -1.8 | (-2.6, -1) | <0.001 |
| 43 | -2.3 | (-3.1, -1.4) | <0.001 |
| 44 | -2.7 | (-3.6, -1.9) | <0.001 |
| 45 | -3.2 | (-4.1, -2.3) | <0.001 |
| 46 | -3.7 | (-4.6, -2.7) | <0.001 |
| 47 | -4.2 | (-5.2, -3.1) | <0.001 |
| 48 | -4.6 | (-5.7, -3.6) | <0.001 |
| 49 | -5.1 | (-6.2, -4) | <0.001 |
| 50 | -5.5 | (-6.6, -4.4) | <0.001 |
| 51 | -6.0 | (-7.1, -4.8) | <0.001 |
| 52 | -6.4 | (-7.5, -5.2) | <0.001 |
| 53 | -6.8 | (-7.9, -5.6) | <0.001 |
| 54 | -7.2 | (-8.3, -6) | <0.001 |
| 55 | -7.5 | (-8.8, -6.3) | <0.001 |
| 56 | -7.9 | (-9.3, -6.6) | <0.001 |
| 57 | -8.3 | (-9.8, -6.8) | <0.001 |
| 58 | -8.7 | (-10.3, -7) | <0.001 |
| 59 | -9.0 | (-10.8, -7.2) | <0.001 |
| 60 | -9.4 | (-11.4, -7.4) | <0.001 |
| 61 | -9.7 | (-12, -7.5) | <0.001 |
| 62 | -10.1 | (-12.6, -7.6) | <0.001 |
| 63 | -10.5 | (-13.2, -7.8) | <0.001 |

**Supplemental Table 4.** Model estimates of Total BabyOSCAR score differences in children with CP GMFCS levels III-V relative to children with CP GMFCS levels I-II, by gestational age (weeks)

| Gestational age (weeks) | Estimate | 95% Confidence intervals | p-value |
| --- | --- | --- | --- |
| 27 | -2.8 | (-7.3, 1.6) | 0.694 |
| 28 | -2.3 | (-6.1, 1.5) | 0.737 |
| 29 | -1.8 | (-5, 1.5) | 0.798 |
| 30 | -1.3 | (-4, 1.5) | 0.880 |
| 31 | -0.8 | (-3, 1.5) | 0.963 |
| 32 | -0.3 | (-2.3, 1.7) | 0.999 |
| 33 | 0.2 | (-1.7, 2) | 1.000 |
| 34 | 0.6 | (-1.3, 2.5) | 0.974 |
| 35 | 0.9 | (-1.2, 2.9) | 0.908 |
| 36 | 1.1 | (-1.1, 3.3) | 0.859 |
| 37 | 1.2 | (-1.2, 3.5) | 0.853 |
| 38 | 1.1 | (-1.3, 3.6) | 0.885 |
| 39 | 0.9 | (-1.5, 3.4) | 0.940 |
| 40 | 0.6 | (-1.8, 3) | 0.987 |
| 41 | 0.2 | (-2.2, 2.6) | 1.000 |
| 42 | -0.4 | (-2.7, 2) | 0.998 |
| 43 | -0.9 | (-3.4, 1.5) | 0.934 |
| 44 | -1.6 | (-4.1, 1) | 0.717 |
| 45 | -2.2 | (-4.9, 0.5) | 0.454 |
| 46 | -2.9 | (-5.7, 0) | 0.261 |
| 47 | -3.5 | (-6.5, -0.5) | 0.145 |
| 48 | -4.1 | (-7.3, -0.9) | 0.079 |
| 49 | -4.6 | (-7.9, -1.3) | 0.040 |
| 50 | -5.1 | (-8.4, -1.8) | 0.019 |
| 51 | -5.5 | (-8.8, -2.2) | 0.009 |
| 52 | -5.8 | (-9.1, -2.6) | 0.004 |
| 53 | -6.1 | (-9.4, -2.8) | 0.002 |
| 54 | -6.4 | (-9.8, -3) | 0.002 |
| 55 | -6.6 | (-10.2, -3.1) | 0.002 |
| 56 | -6.9 | (-10.7, -3) | 0.004 |
| 57 | -7.1 | (-11.3, -2.8) | 0.009 |
| 58 | -7.2 | (-11.9, -2.5) | 0.021 |
| 59 | -7.4 | (-12.7, -2.1) | 0.044 |
| 60 | -7.5 | (-13.4, -1.6) | 0.081 |
| 61 | -7.7 | (-14.3, -1.1) | 0.132 |
| 62 | -7.9 | (-15.1, -0.6) | 0.190 |
| 63 | -8.0 | (-15.9, -0.1) | 0.254 |

**Supplemental Table 5.** Model estimates of BabyOSCAR absolute asymmetry score differences in children with unilateral CP relative to children without CP by gestational age (weeks)

| Gestational age (weeks) | Estimate | 95% Confidence intervals | p-value |
| --- | --- | --- | --- |
| 27 | 1.7 | (0.3, 3) | 0.111 |
| 28 | 1.4 | (0.2, 2.6) | 0.148 |
| 29 | 1.1 | (0.1, 2.1) | 0.212 |
| 30 | 0.8 | (-0.1, 1.6) | 0.349 |
| 31 | 0.5 | (-0.2, 1.2) | 0.617 |
| 32 | 0.2 | (-0.4, 0.8) | 0.938 |
| 33 | 0.0 | (-0.5, 0.5) | 1 |
| 34 | -0.2 | (-0.7, 0.3) | 0.904 |
| 35 | -0.4 | (-0.9, 0.2) | 0.62 |
| 36 | -0.5 | (-1, 0.1) | 0.495 |
| 37 | -0.5 | (-1.1, 0.1) | 0.55 |
| 38 | -0.4 | (-1, 0.3) | 0.76 |
| 39 | -0.2 | (-0.8, 0.4) | 0.978 |
| 40 | 0.1 | (-0.5, 0.7) | 0.997 |
| 41 | 0.4 | (-0.1, 1) | 0.542 |
| 42 | 0.9 | (0.3, 1.4) | 0.029 |
| 43 | 1.3 | (0.7, 1.9) | <0.001 |
| 44 | 1.8 | (1.2, 2.4) | <0.001 |
| 45 | 2.2 | (1.6, 2.9) | <0.001 |
| 46 | 2.7 | (2, 3.4) | <0.001 |
| 47 | 3.2 | (2.4, 3.9) | <0.001 |
| 48 | 3.6 | (2.8, 4.3) | <0.001 |
| 49 | 3.9 | (3.1, 4.7) | <0.001 |
| 50 | 4.2 | (3.4, 5) | <0.001 |
| 51 | 4.5 | (3.6, 5.3) | <0.001 |
| 52 | 4.7 | (3.9, 5.5) | <0.001 |
| 53 | 4.9 | (4, 5.7) | <0.001 |
| 54 | 5.0 | (4.1, 5.9) | <0.001 |
| 55 | 5.1 | (4.2, 6.1) | <0.001 |
| 56 | 5.2 | (4.2, 6.2) | <0.001 |
| 57 | 5.3 | (4.2, 6.4) | <0.001 |
| 58 | 5.4 | (4.1, 6.6) | <0.001 |
| 59 | 5.4 | (4, 6.8) | <0.001 |
| 60 | 5.5 | (3.8, 7.1) | <0.001 |
| 61 | 5.5 | (3.7, 7.3) | <0.001 |
| 62 | 5.6 | (3.6, 7.5) | <0.001 |
| 63 | 5.6 | (3.4, 7.8) | <0.001 |

**Supplemental Table 6.** Model estimates of BabyOSCAR absolute asymmetry score differences among 6 children with unilateral CP relative to 6 children with bilateral CP by gestational age (weeks)

| Gestational age (weeks) | Estimate | 95% Confidence intervals | p-value |
| --- | --- | --- | --- |
| 27 | 1.7 | (0.1, 3.4) | 0.229 |
| 28 | 1.4 | (0, 2.8) | 0.3 |
| 29 | 1.0 | (-0.2, 2.2) | 0.426 |
| 30 | 0.7 | (-0.3, 1.7) | 0.642 |
| 31 | 0.4 | (-0.5, 1.2) | 0.914 |
| 32 | 0.0 | (-0.7, 0.7) | 1 |
| 33 | -0.3 | (-0.9, 0.4) | 0.928 |
| 34 | -0.5 | (-1.1, 0.1) | 0.52 |
| 35 | -0.7 | (-1.4, 0) | 0.265 |
| 36 | -0.8 | (-1.6, -0.1) | 0.195 |
| 37 | -0.8 | (-1.6, -0.1) | 0.213 |
| 38 | -0.8 | (-1.6, 0) | 0.314 |
| 39 | -0.6 | (-1.4, 0.2) | 0.552 |
| 40 | -0.4 | (-1.2, 0.4) | 0.892 |
| 41 | 0.0 | (-0.8, 0.7) | 1 |
| 42 | 0.4 | (-0.4, 1.1) | 0.868 |
| 43 | 0.8 | (0, 1.6) | 0.23 |
| 44 | 1.3 | (0.5, 2.1) | 0.018 |
| 45 | 1.7 | (0.9, 2.6) | <0.001 |
| 46 | 2.2 | (1.2, 3.1) | <0.001 |
| 47 | 2.6 | (1.6, 3.6) | <0.001 |
| 48 | 3.0 | (2, 4.1) | <0.001 |
| 49 | 3.3 | (2.3, 4.4) | <0.001 |
| 50 | 3.6 | (2.5, 4.7) | <0.001 |
| 51 | 3.9 | (2.8, 4.9) | <0.001 |
| 52 | 4.0 | (3, 5.1) | <0.001 |
| 53 | 4.2 | (3.1, 5.3) | <0.001 |
| 54 | 4.3 | (3.2, 5.4) | <0.001 |
| 55 | 4.4 | (3.2, 5.6) | <0.001 |
| 56 | 4.5 | (3.2, 5.7) | <0.001 |
| 57 | 4.5 | (3.1, 5.9) | <0.001 |
| 58 | 4.5 | (2.9, 6.1) | <0.001 |
| 59 | 4.5 | (2.7, 6.4) | <0.001 |
| 60 | 4.6 | (2.5, 6.6) | <0.001 |
| 61 | 4.6 | (2.3, 6.9) | <0.001 |
| 62 | 4.6 | (2, 7.1) | 0.004 |
| 63 | 4.6 | (1.8, 7.4) | 0.012 |

